# Factors Influencing Early Childhood Caries in a University-Based Infant Oral Health Clinic

**DOI:** 10.1101/2025.01.31.25321458

**Authors:** Anum Ijaz, Leda Regina F Mugayar, Khatija Noorullah, Kelly Fernanda Molena, Majd AlSaleh, Sobia Bilal

## Abstract

**Introduction:** Early Childhood Caries (ECC) is a major public health issue that poses significant challenges in pediatric dentistry, affecting infants and preschool children globally. Socioeconomic factors, dietary habits, and parental health literacy play crucial roles in ECC’s development and progression.

**Objective:** The study aims to identify and evaluate factors contributing to ECC in pediatric patients at a university-based infant oral health clinic, with a secondary objective of assessing the effectiveness of newly developed oral health promotional materials through a pilot study.

**Methods:** A retrospective and cross-sectional design was employed to analyze data from 514 pediatric patients. The study examined demographic information, parental and environmental factors, dietary habits, and feeding practices. Statistical analysis was conducted to identify significant predictors of ECC. The pilot study involved 10 participants who provided feedback on the content and face validity of promotional materials.

**Results:** Key factors associated with ECC included race, socioeconomic status, parental health literacy, dietary habits, and bottle use at night. Early detection and preventive strategies, such as regular dental checkups, were found to be critical in reducing caries risk. The pilot study showed positive feedback on promotional materials, indicating relevance, clarity, and visual appeal, with ongoing feedback used to refine the materials.

**Conclusion:** The study highlights the importance of comprehensive preventive strategies that encompass both individual and community-level interventions. It underscores the role of early detection, preventive measures, and tailored oral health education in reducing ECC risk. Limitations include the retrospective design and potential generalization issues due to the study’s focus on a single clinic. Future research should aim to address these limitations through longitudinal studies and larger, more diverse samples to validate the findings and enhance the effectiveness of oral health promotional materials.

## Introduction

Early childhood caries (ECC) is a major public health concern, posing a persistent challenge in pediatric dentistry not only in the United States but globally. This condition affects a significant proportion of infants and preschool children and is particularly prevalent among socioeconomically disadvantaged populations (Anil & Anand, 2017). The American Academy of Pediatric Dentistry (AAPD) defines ECC as the presence of one or more decayed, missing (due to decay), or filled tooth surfaces in any primary tooth of a child under six years of age. ECC tends to progress quickly, driven by factors like high sugar consumption, inadequate oral hygiene, and a lack of early dental preventive care. This rapid progression is especially common in high-risk children and is often inadequately addressed, leading to severe consequences such as dental pain, tooth loss, and broader impacts on physical, psychological, and social well-being. These repercussions can influence a child’s nutritional intake, speech, self-esteem, and overall quality of life (Swamy et al., 2012; Kotha et al., 2022).

ECC has far-reaching effects on a child’s overall health, growth, and development, influencing quality of life for both children and their families. Beyond impairing oral health, it can lead to serious physical, mental, and social issues. In the short term, untreated ECC causes pain, infection, disrupted sleep, poor appetite, hospitalizations, school absences, learning difficulties, reduced concentration, and premature tooth loss, which can result in chewing problems. Long-term consequences include poor oral health, a higher risk of caries in permanent teeth, stunted physical growth, and psychological challenges, all impacting quality of life (Mansoori et al., 2019). In addition, the financial burden of treating ECC also places stress on families and healthcare systems, underscoring the need for preventive measures, ideally initiated during the child’s first dental visit, recommended within 6 months of the first primary tooth eruption and not later than 12 months (Prakash et al., 2006). Notably, recent research suggests potential links between untreated ECC and adverse cognitive and neurodevelopmental outcomes in young children, as this disease often occurs during a critical period of brain development (Foláyan et al., 2023).

A comprehensive review of the literature reveals multiple risk factors contributing to the development of ECC. Significant maternal health factors include gestational diabetes and smoking during pregnancy, both recognized as potent predictors of ECC. Additionally, the mode of delivery influences its risk, with cesarean sections associated with higher rates of ECC due to altered patterns of bacterial colonization. Although breastfeeding is generally promoted for its health benefits, prolonged and nocturnal breastfeeding has been linked to an increased risk of ECC, underscoring the necessity for dental care education among breastfeeding mothers. Furthermore, parental perceptions and knowledge about oral health profoundly impact the effectiveness of ECC prevention practices. Environmental tobacco smoke exposure during early childhood also correlates directly with elevated ECC risk, highlighting the complex, multifactorial nature of ECC (Nakayama & Mori, 2017; Bernabé et al., 2017).

Despite the recognition of these risk factors and the widespread adoption of preventive measures, dental caries remains a persistent issue among pediatric populations. A systematic review spanning a decade found an average ECC prevalence of 23.8% among children under 36 months and 57.3% among children aged 36 to 71 months across multiple countries (Tantawi et al., 2018). In developing countries like India, ECC is a significant issue, with a reported prevalence of about 49.6% (Ganesh et al., 2018). A comprehensive study using WHO diagnostic criteria reported ECC prevalence in the Americas at 48%, indicating that nearly half of preschool-age children in this region are affected (Uribe et al., 2021), which highlights the public health impact. The United States has also seen an unsettling trend in ECC prevalence. Between 2013 and 2018, the rates initially decreased from 19.6% to 17.4%, but then rose again to 18.7%. Severe Early Childhood Caries (S-ECC) prevalence increased from 9.8% to 11.9%, suggesting a worsening situation among affected children (Kotha et al., 2022). Data from the 2015–2016 National Health and Nutrition Examination Survey (NHANES) shows an increase in prevalence across age groups, from 21.4% in the 2–5 age group to over 50% in older children, indicating the progression of untreated caries (Fleming & Affoul, 2018).

This persistence in prevalence indicates a gap in the effectiveness of current preventive strategies, possibly due to challenges in their universal implementation and accessibility across various socio-economic groups. Current preventive strategies, including fluoride varnish applications, oral health education for parents during prenatal visits, and early dental check-ups by the age of one year, often encounter limitations. These include insufficient reach within underserved populations and a lack of adherence to prevention protocols by caregivers. The effectiveness of these interventions is further diminished by social determinants such as socioeconomic status and access to healthcare. These challenges underscore the need for more inclusive and community-specific health promotion strategies that are sensitive to the socio-economic and cultural contexts of targeted populations (Majorana et al., 2014).

This study aims to rigorously assess the factors influencing pediatric patients’ caries experience, beginning with a comprehensive review within a university-based infant oral health clinic. By examining referral patterns, pregnancy and birth-related factors, dietary habits, parental dental history, and perceptions of general and oral well-being, this study will delineate the complex genetic, environmental, and behavioral contributors to ECC risk.

Specifically, this study will explore:

- The influence of referral and perinatal factors on ECC.
- Dietary patterns and their relationship with ECC.
- The impact of parental dental history and perceptions on ECC prevention.
- The association of household tobacco use with children’s oral health.
- How maternal health conditions during pregnancy and the mode of delivery may affect children’s susceptibility to ECC.
- The connection between children’s overall health and dental conditions.

Secondary objectives include developing and adapting oral health promotion materials based on the insights gained. This involves piloting new educational resources and evaluating their effectiveness through both face and content validity assessments, aiming to enhance preventive efforts and improve oral health outcomes across diverse pediatric care settings.

This comprehensive approach seeks to address existing knowledge gaps and leverage this information to refine and enhance oral health strategies tailored not only to children attending specialized clinics but also to broader pediatric populations.

## Materials and Methods

### Study Design

This study employs a retrospective and cross-sectional design to explore the factors associated with Early Childhood Caries (ECC) in pediatric patients attending a university-based infant oral health clinic. The retrospective component involves the examination of existing patient records to assess historical data, while the cross-sectional aspect allows for the analysis of data at a single point in time. This method is particularly employed to meet the secondary objectives of the study, such as evaluating and enhancing oral health promotion materials based on contemporary data insights.

### Study Setting and Population

The research is conducted at the Infant Oral Health Clinic at the University of Illinois Chicago (UIC) College of Dentistry, which caters to a diverse pediatric demographic. As a primary care facility, it focuses on the oral health of infants and children up to the age of six. The initial dataset included records for 801 pediatric patients. After rigorous data cleaning to ensure accuracy and reliability, 287 records were excluded due to incomplete data, resulting in a robust final dataset of 514 patients. This varied sample is critical for understanding the multifaceted influences on ECC across different demographic and socio-economic groups.

### Data Collection Methods

Data collection is conducted using the axiUm software system, specifically tailored for use in dental clinics. This process, active from September 2020 to August 2023, captures a detailed snapshot of pediatric patients’ experiences, ensuring a comprehensive representation of the patient cohort over this period. The types of data collected include medical and dental histories, Infant Caries Risk Assessment Forms, Infant Medical History Forms, and additional relevant documentation that provides comprehensive insights into each patient’s health background and risk factors for ECC. Variables such as referral sources, pregnancy and birth-related factors, feeding practices, parental health history, and the presence of tobacco smoke exposure are meticulously recorded.

As part of the secondary objectives, a pilot study is being conducted to gather data on the content and face validity of both modified and newly developed materials (Appendix 1). These materials have been designed based on certain findings from the primary objectives of the study. Participants were presented with the modified promotional material in English and Spanish and asked to provide their feedback. Responses were obtained through a structured questionnaire consisting of five questions. Participants responded to each question with either yes/no or by rating on a scale from extremely useful/helpful to not at all.

### Data Analysis Plan

The collected data has been analyzed using SPSS software to perform statistical tests that assess relationships between the identified risk factors and the incidence of ECC. Descriptive statistics was used to summarize the data, while inferential statistics, including logistic regression models, was utilized to explore associations between independent variables (such as maternal health during pregnancy, mode of delivery, and feeding practices) and the dependent variable (presence of ECC). The analysis will also include tests for statistical significance and confidence intervals to determine the robustness of the associations.

This methodological approach ensures a thorough examination of the factors influencing ECC in a well-defined pediatric population, enabling the identification of targeted interventions for improving oral health outcomes. Furthermore, regarding secondary objectives, responses to the questionnaire were analyzed to determine the percentage of participants who answered affirmatively to each question.

## Results

Table 1 shows the baseline characteristics of the study subjects, where 352 out of 514 pediatric patients reported caries, resulting in a caries rate of 68.5%. It covers a range of factors, including demographics, parental-related factors, environmental factors, child-related factors, and feeding and nutrition-related factors. Significant variables identified in the table include race, with White patients exhibiting a higher caries rate (72.3%) compared to Black and Asian patients. The referral source also showed significance, with most carious patients referred by dentists (88.9%, p < 0.001). The child’s age was also significant (p < 0.001), with age 3 having the highest caries rate (87.1%). The presence of obvious plaque and/or bleeding gums was strongly associated with caries (89.7%, p < 0.001), highlighting the importance of oral hygiene. In terms of feeding patterns, current bottle use (61.8%, p = 0.008) and bottle use at night (60.4%, p = 0.013) were both significantly associated with caries. Longer breastfeeding durations were also significant (p = 0.002), indicating potential feeding-related factors in caries prevalence.

**Table 1:**
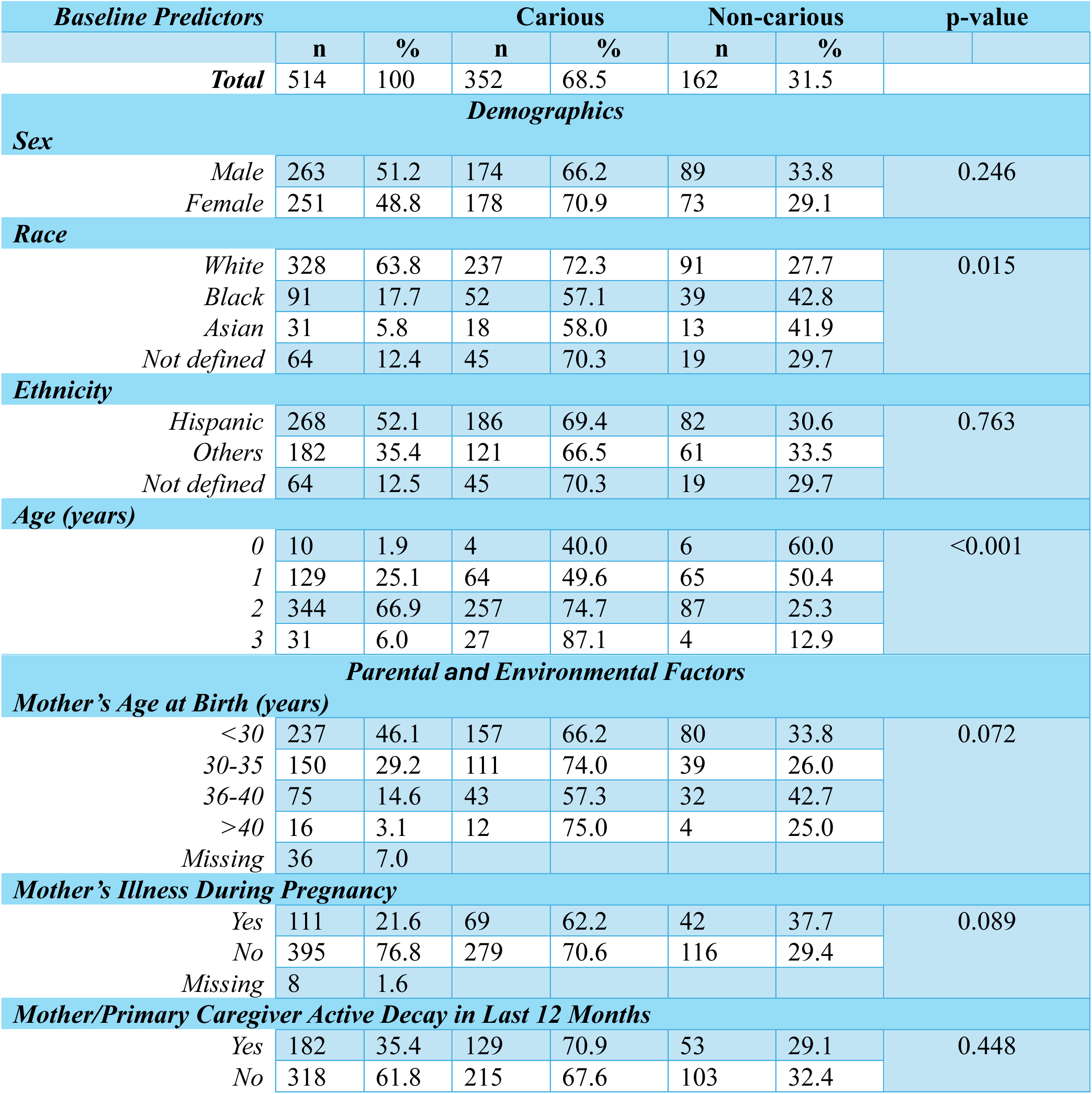
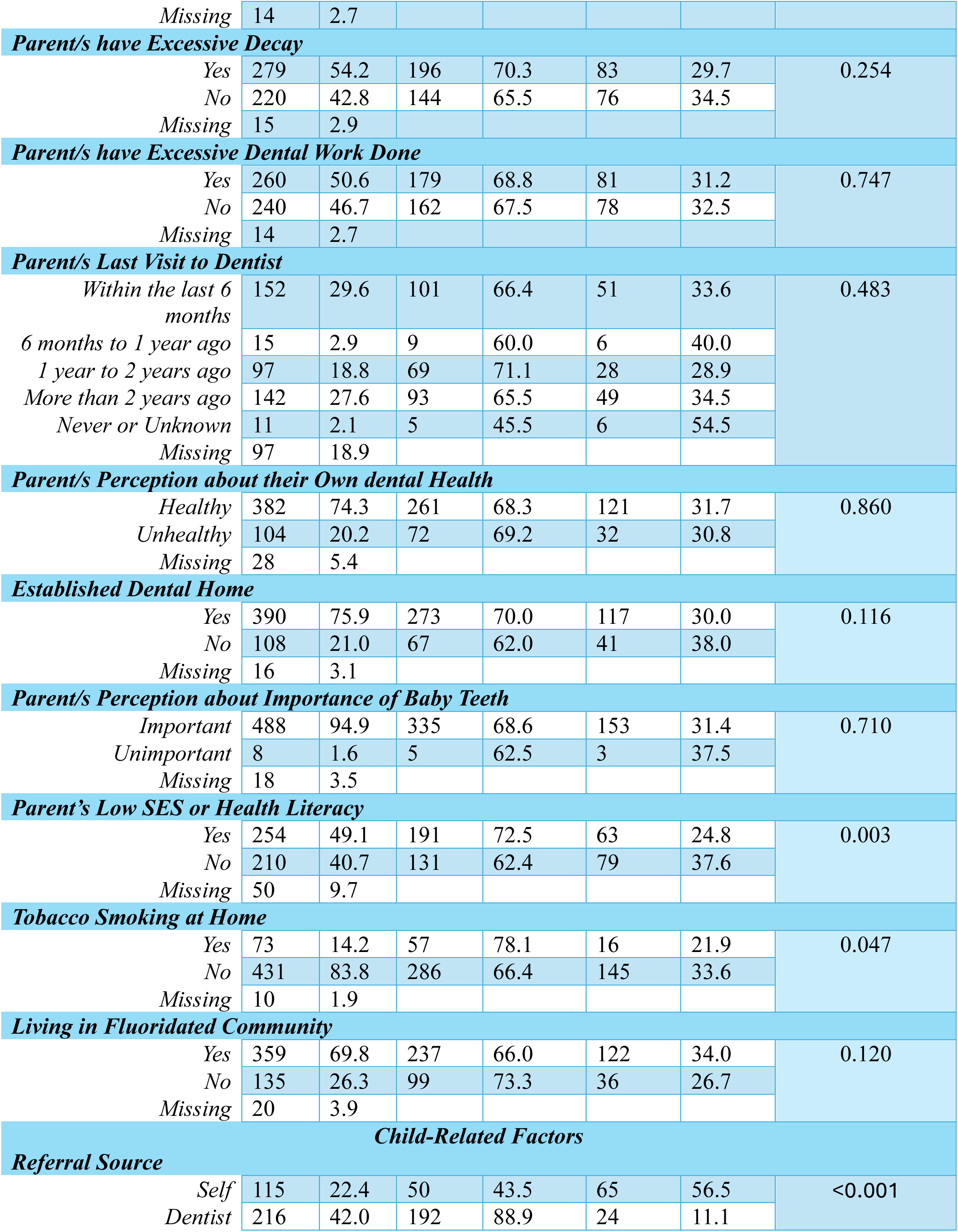
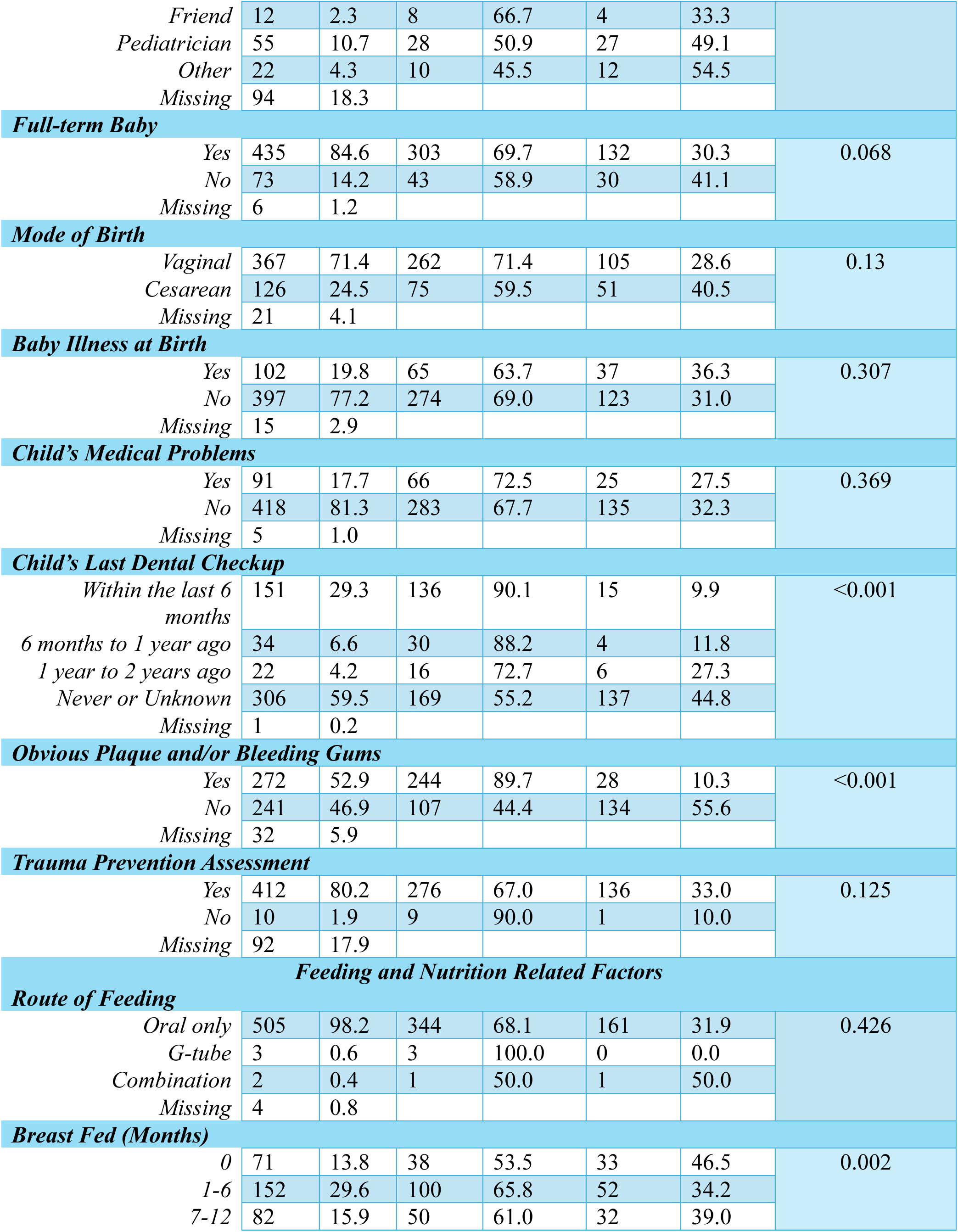
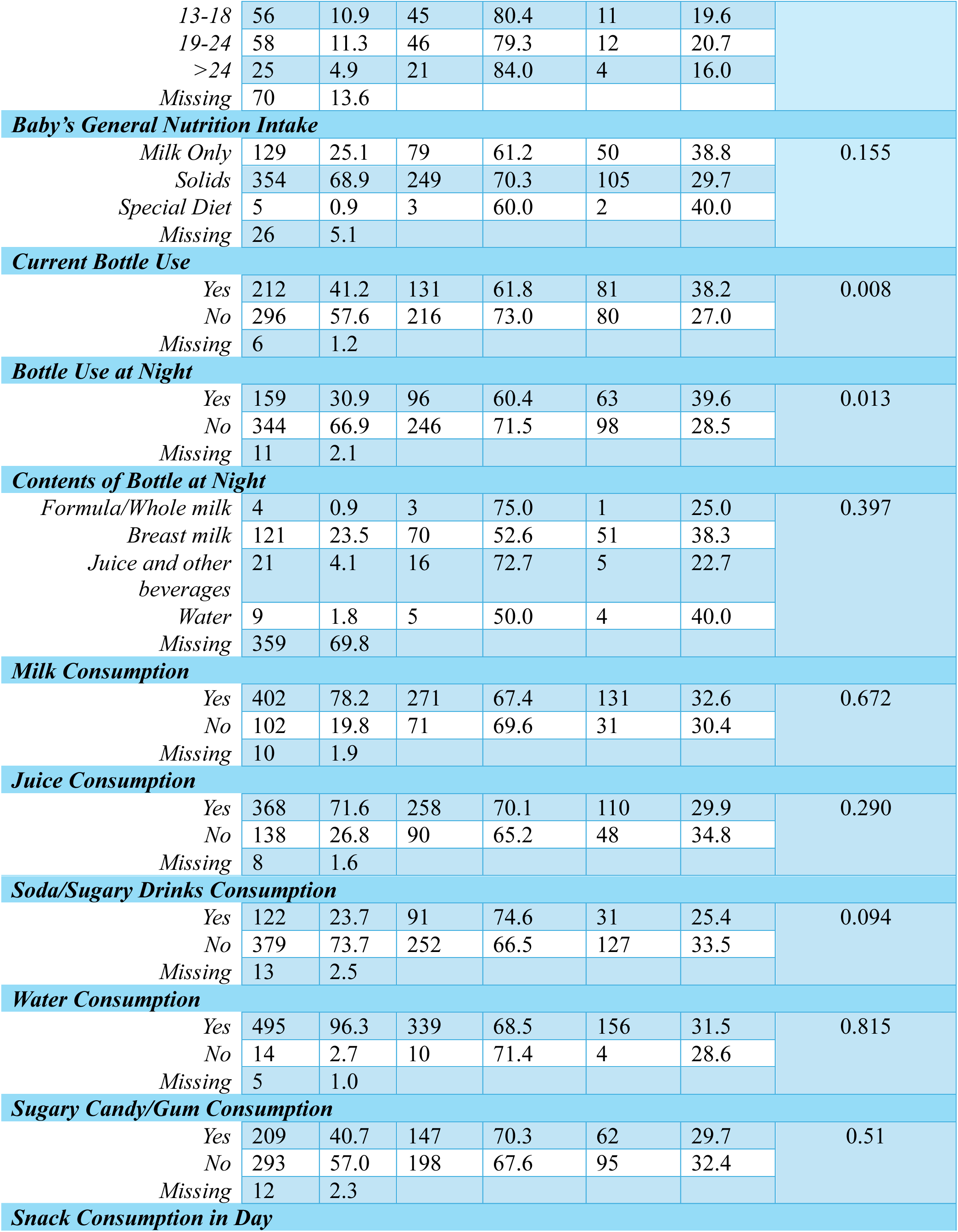
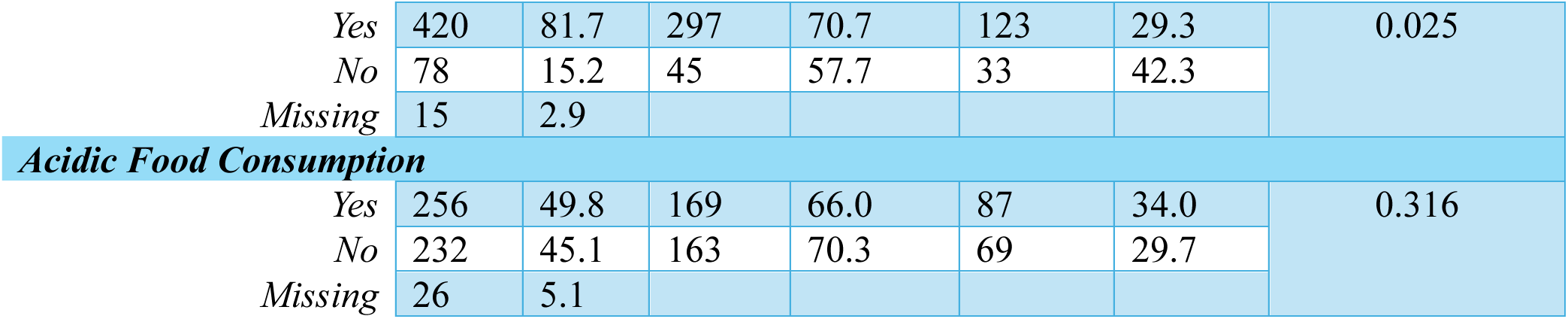
Baseline Characteristics and Predictors of Early Childhood Caries.

Table 2 displays univariate analysis with crude odds ratios (OR) and multivariate analysis with adjusted ORs for pediatric caries, including corresponding confidence intervals (CI) and p-values to highlight significant factors. Regarding race, Black children showed a crude OR of 1.95 (95% CI: 1.21 - 3.16, p-value 0.006) for caries as compared to White children, but it decreased to an adjusted OR of 1.74 (95% CI: 0.77 - 3.89, p-value 0.181) and became non-significance after adjustment. Concerning age, the crude ORs for ages 1 and 2 compared to age 3 are 0.23 (95% CI: 0.06 - 0.82, p-value 0.024) and 0.09 (95% CI: 0.02 - 0.51, p-value 0.006), respectively. These associations also become non-significant upon multivariate analysis, with an OR of 3.46 (95% CI: 0.85 - 14.06, p-value 0.083) for age 1, and 1.52 (95% CI: 0.39 - 5.95, p-value 0.548) for age

**Table 2:** Univariate and Multivariate Analysis of Factors Associated with Early Childhood Caries.

| *Variables* | **Crude OR**<br>**(95% CI)** | **p-value** | **Adjusted OR**<br>**(95% CI)** | **p-value** |
| --- | --- | --- | --- | --- |
| ***Race*** |  |  |  |  |
| *White* | 1.00 (Ref) |  |  |  |
| *Black* | 1.95 (1.21, 3.16) | 0.006 | 1.74 (0.77,3.89) | 0.181 |
| *Asian* | 1.88 (0.89, 3.99) | 0.100 | 0.65 (0.19, 2.15) | 0.484 |
| *Not defined* | 1.10 (0.61, 1.98) | 0.752 | 0.52 (0.20, 1.32) | 0.170 |
| ***Age (Years)*** |  |  |  |  |
| *0* | 0.68 (0.18, 2.51) | 0.560 | 2.38 (0.19,28.52) | 0.493 |
| *1* | 0.23 (0.06, 0.82) | 0.024 | 3.46 (0.85,14.06) | 0.083 |
| *2* | 0.09 (0.02, 0.51) | 0.006 | 1.52 (0.39,5.95) | 0.548 |
| *3* | 1.00 (Ref) |  |  |  |
| ***Referral Source*** |  |  |  |  |
| *Self* | 1.00 (Ref) |  |  |  |
| *Dentist* | 0.09 (0.06, 0.17) | <0.001 | 0.12 (0.05, 0.29) | <0.001 |
| *Friend* | 0.39 (0.11, 1.35) | 0.136 | 0.58 (0.11, 3.04) | 0.520 |
| *Pediatrician* | 0.74 (0.39, 1.41) | 0.364 | 0.61 (0.24, 1.52) | 0.286 |
| *Other* | 0.92 (0.37, 2.31) | 0.864 | 0.59 (0.15, 2.36) | 0.463 |

| ***Parent's Low SES or Health Literacy*** |  |  |  |  |
| --- | --- | --- | --- | --- |
| Yes | 1.00 (Ref) |  |  |  |
| No | 1.83 (1.23, 2.72) | 0.003 | 1.13 (0.59, 2.13) | 0.717 |
| ***Tobacco Smoking at Home*** |  |  |  |  |
| Yes | 1.00 (Ref) |  |  |  |
| No | 1.81 (1.00, 3.26) | 0.049 | 0.76 (0.31, 1.90) | 0.563 |
| ***Child's Last Dental Checkup*** |  |  |  |  |
| Within the last 6 months | 1.14 (0.08, 0.24) | <0.001 | 0.15 (0.07, 0.34) | <0.001 |
| 6 months to 1 year ago | 0.16 (0.06, 0.48) | <0.001 | 0.19 (0.05, 0.76) | 0.019 |
| 1 year to 2 years ago | 0.46 (0.18, 1.21) | 0.117 | 0.84 (0.20, 3.47) | 0.81 |
| Never or Unknown | 1.00 (Ref) |  |  |  |
| ***Obvious Plaque and/or Bleeding Gums*** |  |  |  |  |
| Yes | 1.00 (Ref) |  |  |  |
| No | 10.91 (6.85, 17.39) | <0.001 | 9.12 (4.63, 17.98) | <0.001 |
| ***Breast Fed (Months)*** |  |  |  |  |
| 0 | 1.00 (Ref) |  |  |  |
| 1-6 | 0.59 (0.34, 1.06) | 0.080 | 0.62 (0.25, 1.52) | 0.293 |
| 7-12 | 0.74 (0.39, 1.40) | 0.353 | 0.33 (0.12, 0.93) | 0.037 |
| 13-18 | 0.28 (0.13, 0.63) | 0.002 | 0.12 (0.04, 0.41) | <0.001 |
| 19-24 | 0.30 (0.14, 0.66) | 0.003 | 0.18 (0.05, 0.63) | 0.007 |
| >24 | 0.22 (0.07, 0.70) | 0.011 | 0.37 (0.07, 2.03) | 0.255 |
| ***Current Bottle Use*** |  |  |  |  |
| Yes | 1.00 (Ref) |  |  |  |
| No | 0.59 (0.41, 0.87) | 0.008 | 0.87 (0.32, 2.36) | 0.783 |
| ***Bottle Use at Night*** |  |  |  |  |
| Yes | 1.00 (Ref) |  |  |  |
| No | 0.61 (0.41, 0.90) | 0.013 | 1.18 (0.42, 3.33) | 0.757 |
| ***Snack Consumption in Day*** |  |  |  |  |
| Yes | 1.00 (Ref) |  |  |  |
| No | 1.77 (1.08, 2.91) | 0.024 | 1.06 (0.48, 2.32) | 0.889 |

For referral source, being referred by a dentist has a crude OR of 0.09 (95% CI: 0.06 - 0.17, p-value <0.001), with an adjusted OR of 0.12 (95% CI: 0.05 - 0.29, p-value <0.001), indicating lower caries risk. Low socioeconomic status (SES) or health literacy had a crude OR of 1.83 (95% CI: 1.23 - 2.72, p-value 0.003), which reduced and became insignificant to 1.13 (95% CI: 0.59 - 2.13, p-value 0.717) on adjusting with other variables.

For oral health indicators, plaque or bleeding gums initially showed a significant crude OR of 10.91 (95% CI: 6.85 - 17.39, p < 0.001), which remained significant after adjustment, with an OR of 9.12 (95% CI: 4.63 - 17.98, p < 0.001). Dental checkups within the last 6 months or between 6 months to 1 year ago initially showed significant crude ORs of 0.14 (95% CI: 0.08 - 0.24, p < 0.001) and 0.16 (95% CI: 0.06 - 0.48, p < 0.001) respectively. After multivariate analysis, the ORs were 0.15 (95% CI: 0.07 - 0.34, p < 0.001) and 0.19 (95% CI: 0.05 - 0.76, p = 0.019) respectively, remaining still significant.

Furthermore, tobacco smoking at home initially demonstrated a crude OR of 1.81 (95% CI: 1.00 - 3.26, p = 0.049), which decreased to 0.76 (95% CI: 0.31 - 1.90, p = 0.563) after considering other variables, indicating non-significance. Extended breastfeeding for 13-18 months and 19-24 months had adjusted ORs of 0.12 (95% CI: 0.04 - 0.41, p < 0.001) and 0.18 (95% CI: 0.05 - 0.63, p = 0.007) respectively, suggesting lower caries risk. However, breastfeeding beyond 24 months showed no significant benefits, with an OR of 0.37 (95% CI: 0.07 - 2.03, p = 0.255). Similarly, bottle use at night initially exhibited a crude OR of 0.61 (95% CI: 0.41 - 0.90, p = 0.013) and adjusted OR of 1.18 (95% CI: 0.42 - 3.33, p = 0.757), indicating a non-significant association after considering other variables. Snack consumption during the day initially showed a crude OR of 1.77 (95% CI: 1.08 - 2.91, p = 0.024), which lost significance in the adjusted analysis with an OR of 1.06 (95% CI: 0.48 - 2.32, p = 0.889).

Results from the pilot study evaluating oral health promotional materials currently involved 10 participants to gauge content and face validity, displayed in figure 1. The majority of participants found the materials relevant, clear, visually appealing, and useful. In terms of relevance and importance, 90% of participants indicated that the flyer addressed topics significant to them and relevant to their health concerns or interests. Regarding clarity and comprehension, 100% found the information easy to understand. Similarly, the visual appeal of the flyer was well received, with all participants indicating that it caught their eye and encouraged them to read further. The balance of text and pictures was also satisfactory, with all participants indicating that they did not prefer more pictures over text. Furthermore, in terms of usefulness and call to action, 100% of participants stated that the flyer was extremely useful, providing clear guidance on what actions to take.

**Fig 1:**
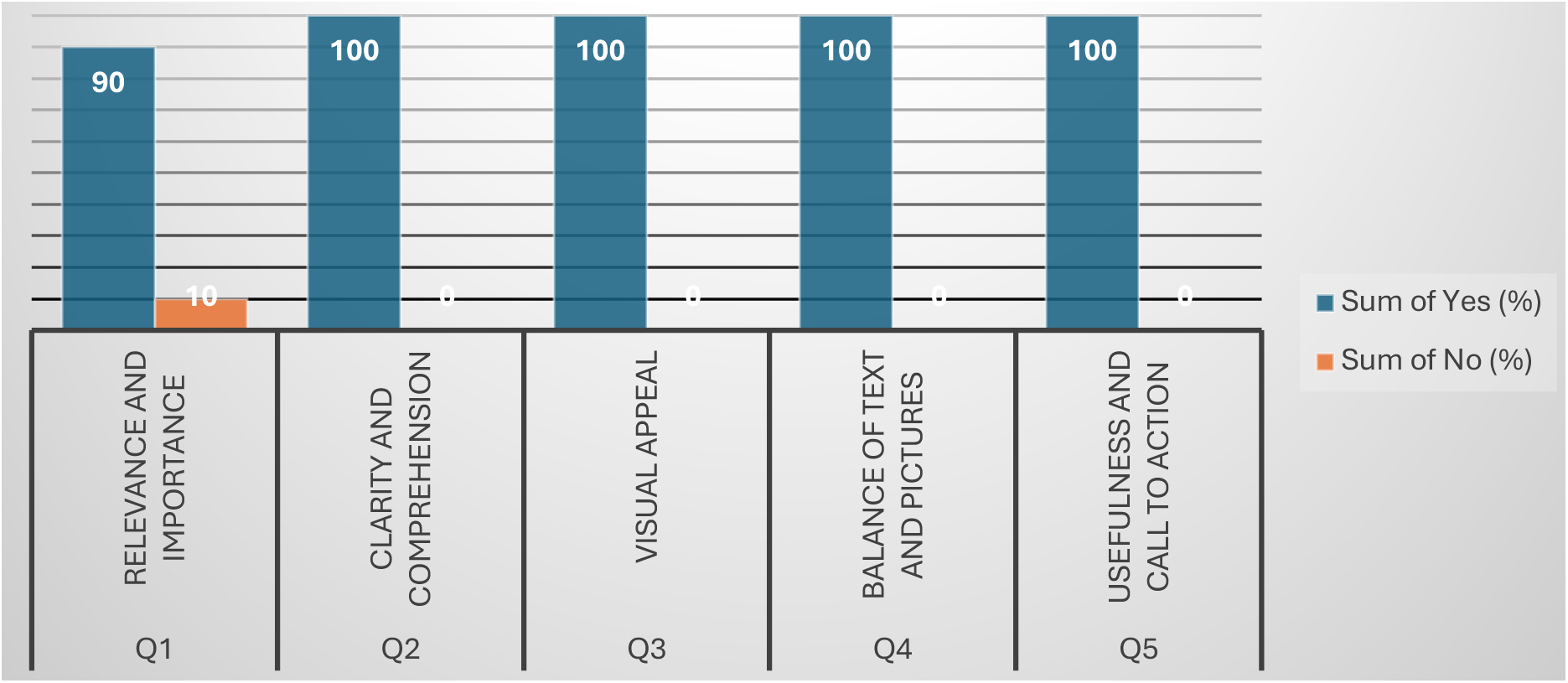
Oral Health Promotion Material: Pilot Study Feedback.

## Discussion

### Demographic Factors

Race has long been considered a significant factor in ECC. Previous studies have identified a complex relationship between race/ethnicity and ECC prevalence, with non-White racial groups such as Black, Hispanic, and Asian children having higher odds of developing ECC compared to their White counterparts (Psoter et al., 2006; Shiboski et al., 2003). This racial disparity has often been attributed to socioeconomic factors such as education, income, and access to affordable healthcare. In our study, Black children were initially 95% more likely to experience ECC compared to White children. However, after adjusting for other socioeconomic factors— including socioeconomic status (SES), parental health literacy, tobacco smoking, and dietary factors—the significance of race diminished. This suggests that socioeconomic determinants might play a pivotal role in explaining the racial disparities observed in ECC prevalence (Rai & Tiwari, 2018).

These findings indicate that while racial disparities in ECC exist, they may be driven by deeper socioeconomic issues. Low-income individuals and specific racial groups, like Black and Mexican American populations, exhibit a higher prevalence and severity of dental caries (Psoter et al., 2006; Shiboski et al., 2003). Additionally, given that smaller datasets can reduce statistical power, impacting the detection of significant relationships, further research with larger sample sizes is needed to validate these findings and explore these relationships more comprehensively.

### Child-related Factors

The majority of children who presented at UIC Infant Oral Health Clinics were referred by general dentists, a factor associated with significantly lower caries rates. The crude and adjusted odds ratios for fewer caries among these children were 0.09 and 0.12, respectively, indicating that early detection and treatment, along with improved oral care education, are crucial in reducing the incidence of caries. Furthermore, regular dental checkups proved to be highly protective, with children who attended a checkup within the past six months having about 85% lower chances of developing caries. This highlights the importance of consistent dental care, preventive measures, and a holistic approach to managing early childhood caries (ECC) in pediatric populations.

Earlier studies have shown that training pediatricians to detect ECC and refer cases to dentists has been shown to significantly reduce caries incidence. A study found that trained pediatricians demonstrated high accuracy in identifying ECC and referring children for dental care, which plays a crucial role in lowering caries rates (Elhoseny & Elshazly, 2010). Primary care providers, including pediatricians, are central to early detection and referral, with training in oral health promotion leading to better referral rates and improved dental health outcomes (Kagihara et al., 2009). These initiatives can reduce the number of decayed, missing, and filled tooth surfaces in children aged 3 to 4 years (Braun et al., 2017).

Child age also plays a critical role in the risk of ECC. A longitudinal study found that as children grow older, their risk of caries increases, particularly if they exhibit early signs of caries or do not consistently follow preventive measures. This pattern is especially evident among three-year-old, who tend to have the highest caries rates. Contributing factors such as high sugar intake and poor oral hygiene may accelerate the increase in caries over time (Meurman & Pienihäkkinen, 2010). To counter these risks, school-based interventions have shown promise. A randomized trial with children aged 3-5 years demonstrated that consistent preventive measures in school settings can significantly reduce caries incidence. This emphasizes the importance of early preventive strategies, especially for older children who might be at greater risk without consistent application of these strategies. School-based programs, combined with parental guidance and regular dental care, can be crucial in reducing the risk of ECC in children as they grow (Samuel et al., 2019).

Another important factor is the presence of visible plaque or bleeding gums, both of which are strongly associated with a higher likelihood of caries. This further reinforces the need for early oral hygiene practices and parental education. Studies have shown that visible plaque on the labial surfaces of the maxillary incisors is a significant sign of caries risk in children (Alaluusua & Malmivirta, 1994), and research on adults also revealed that dental caries was positively associated with the severity of periodontal disease, suggesting that high plaque scores and bleeding on probing are correlated with increased risk (Durand et al., 2019; Kallio et al., 1997).

### Parental-related and Environmental Factors

Socioeconomic status (SES) and health literacy significantly impact the risk of ECC. In our study, we initially found that low SES or health literacy was associated with an 83% higher chance of caries. However, after adjusting for other variables, this association became non-significant. Mixed results have been observed in the research literature regarding this topic. Some studies indicate a link between low SES, defined by factors such as education, income, and occupation, and a higher incidence of dental caries (Costa et al., 2012). A national survey among Danish adults revealed that individuals with lower education, lower income, and those receiving social benefits were more likely to exhibit inadequate health literacy, suggesting that health literacy could be intertwined with SES and influence health outcomes (Svendsen et al., 2020).

On the other hand, some studies found that low parental oral health literacy seems to affect the occurrence of at least one clinical consequence of untreated dental caries in children, but not necessarily the presence of untreated caries itself (Martins et al., 2021). Another study concluded that maternal education levels were linked to lower rates of early childhood caries, while SES showed no significant influence on these rates (Bhardwaj & Bhardwaj, 2014). These findings suggest that the relationship between SES, health literacy, and ECC is complex, with various factors influencing outcomes. Further research is needed to understand these interactions and their effects on early childhood caries.

Tobacco smoking at home also impacts the risk of ECC. Our study found that children from smoking households had an 81% higher risk of developing caries, but this risk became non-significant after adjusting for other factors, suggesting complex interactions. However, several studies have consistently found a significant association between environmental tobacco smoke (ETS) and dental caries. For instance, one study reported that children from smoking households had a higher prevalence of caries compared to non-smoking households, even after adjusting for factors like age, SES, and toothbrushing frequency (Shenkin et al., 2004). Additionally, a cross-sectional study of preschool children showed that household smoking exposure increased the risk of dental caries, with a dose-response relationship between cumulative smoking and caries (Goto et al., 2019). Furthermore, research into maternal smoking during pregnancy and postnatal household smoking revealed that both prenatal and postnatal exposure to tobacco smoke could elevate the risk of caries in children (Tanaka et al., 2009).

### Dietary Factors

Extended breastfeeding appears to have a protective effect against ECC, with children breastfed for 13-24 months being 82-88% less likely to experience caries. In contrast, breastfeeding beyond 24 months does not show a significant reduction in risk. Several studies suggest that prolonged breastfeeding, particularly beyond 12 months, may be associated with a higher risk of dental caries. A study in southern Brazil showed that children breastfed for 24 months or longer had a higher number of decayed, missing, or filled primary tooth surfaces (dmfs) and a 2.4 times greater risk of severe early childhood caries (S-ECC) compared to those breastfed for up to 12 months (Peres et al., 2017). Similarly, another study indicated that prolonged breastfeeding might increase caries risk, whereas breastfeeding for shorter durations (13-23 months) did not significantly impact it (Kramer et al., 2007). The Generation R Study, a prospective multiethnic cohort in Rotterdam, also found that prolonged breastfeeding (beyond 12 months) and nocturnal bottle-feeding were linked to dental caries, suggesting that feeding frequency and oral hygiene might also play a role (Lunteren et al., 2021). Overall, while breastfeeding for 13-24 months seems protective, prolonged breastfeeding beyond this period may increase caries risk, emphasizing the need for preventive measures.

Snack consumption is also a significant factor in the risk of ECC. Children who snack during the day are about 77% more likely to develop caries than those who do not, highlighting the impact of diet and frequent snacking on dental health. A case-control study in India found that snacking more than three times per day was significantly associated with ECC, with an odds ratio (OR) of 2.78 (Mahesh et al., 2013). Similarly, a study in Seoul, Korea, indicated that a higher frequency of between-meal snacks was linked to a higher prevalence of ECC (Jin et al., 2003). However, multivariate analyses in our study showed that this association might not always be statistically significant, suggesting that frequency of snacking may be more critical than merely recording the consumption of snacks. Additionally, this study focused on hospital-visiting patients and families, which might suggest higher awareness about snacking and better oral health care. Further qualitative research is needed to confirm this hypothesis and to understand the nuances of snacking behavior and its effect on ECC risk.

Furthermore, continual bottle use and bottle use at night initially seemed to increase the risk ofECC, but multivariate analyses suggest that other factors may play a more significant role. A case-control study found that while bottle-feeding itself did not represent a significant risk for ECC, night-time consumption of sweet beverages after the first 24 months significantly increased the risk (Mahesh et al., 2013). Similarly, a study in Croatia revealed that bottle-feeding alone was not associated with ECC risk, but night-time consumption of sweet beverages and lack of early teeth-brushing significantly increased the risk (Lulić-Dukić et al., 2001). These findings suggest that factors like the type of liquid in the bottle and feeding behaviors are more critical in determining ECC risk than the mere act of bottle-feeding. Emphasizing proper feeding practices and transitioning children from bottles to cups at an appropriate age could help reduce the risk of ECC. Parents and caregivers should be advised to avoid sweet beverages in bottles, especially at night, and to ensure early teeth-brushing and regular dental visits to minimize the risk of ECC.

### Impact on Prevention and Intervention Strategies

The findings from our study have potential implications for developing targeted prevention and intervention strategies for managing early childhood caries (ECC). By identifying specific factors that contribute to ECC, such as racial disparities, dietary habits, bottle use, parental-related factors, and environmental factors, these insights can help inform tailored preventive measures.

For instance, our study suggests that parental education and regular dental checkups play critical roles in reducing caries risk. This finding could support the implementation of community-based educational programs that promote oral health and the importance of regular dental visits. Early intervention, particularly with infants and toddlers, can be crucial in managing ECC. Training programs for healthcare providers, such as pediatricians and dentists, can further enhance early detection and appropriate referrals for dental care.

The study’s results also highlight the importance of addressing broader socioeconomic determinants, reinforcing the need for public health policies that promote equitable access to healthcare and educational resources. Moreover, the focus on proper feeding practices, such as transitioning from bottle-feeding to cups at an appropriate age and avoiding sugary beverages at night, can lead to more effective ECC prevention strategies.

Moreover, early childhood caries (ECC) and its associated risk factors should become integral topics during prenatal and neonatal appointments, laying the groundwork for proactive prevention and intervention strategies. Prenatal visits offer a crucial opportunity to educate expectant parents about ECC risks, emphasizing the significance of dietary habits, bottle use, and socioeconomic factors. This early awareness will enable parents to make informed decisions, such as transitioning from bottle-feeding to cups at an appropriate age and avoiding sugary beverages, ultimately influencing infant feeding practices and early dental care.

Neonatal appointments serve as reinforcement, emphasizing the importance of early oral hygiene and regular dental checkups. Healthcare providers can guide parents on effective prevention measures and the role of early intervention in reducing ECC risk. By integrating ECC education into these appointments, a holistic approach to oral health can emerge, empowering parents to prioritize their child’s dental care from the outset. This proactive stance will not only reduce ECC incidence but will also establishes the foundation for healthier dental outcomes, underscoring the notion that good oral health begins before the first tooth appears.

The pilot study is still ongoing to gather more data and results. Based on the findings, the intervention in the form of a communication plan will be developed to ensure effective dissemination of information. The initial steps in this plan focus on refining promotional materials, which involves minor adjustments to maintain a good balance between text and images, ensuring that visuals support key messages without causing distractions. It also aims to cater to a broader audience by providing materials in diverse languages. Currently, materials are available in both English and Spanish, promoting inclusivity and reaching a wider demographic.

For distribution, the plan involves utilizing dental clinics, pediatric care facilities, and community centers to ensure the promotional materials reach the right audience. Collaboration with healthcare providers will further expand the reach, ensuring a comprehensive distribution strategy. To ensure the effectiveness of the communication plan over time, a system for continuous feedback will be implemented. This will allow for ongoing adjustments based on user input, maintaining the relevance and effectiveness of the promotional materials. Lastly, the plan will incorporate education and awareness campaigns, leveraging the promotional materials to raise public awareness about oral health and other relevant topics. These campaigns will emphasize key messages and encourage healthy practices, ultimately contributing to a broader understanding and improvement of public health outcomes.

### Limitations and Future Research

While our study offers valuable insights into ECC risk factors, it has certain limitations. The retrospective and cross-sectional design may restrict the ability to infer causal relationships. Additionally, our focus on a university-based infant oral health clinic could limit the generalizability of the results to other pediatric populations.

Future research could address these limitations by conducting longitudinal studies that track caries development over time. This would provide a clearer understanding of the progression of ECC and the influence of various risk factors. Additionally, exploring the impact of other variables, such as genetic factors and cultural influences, could offer a more comprehensive perspective on ECC.

Qualitative studies could also be valuable in gaining deeper insights into parental attitudes and behaviors regarding oral health. This would help refine educational and preventive strategies to ensure they resonate with diverse communities. Further research on the effectiveness of different intervention programs and public health policies could lead to more robust and impactful approaches to ECC prevention.

Despite the success of the pilot study in gathering initial feedback on promotional materials, its findings might not fully represent a broader audience due to its limited sample size. Further research should focus on refining these promotional materials based on a larger and more diverse participant pool to ensure relevance and effectiveness across different populations.

## Conclusion

Our study identified several significant factors contributing to the development of early childhood caries (ECC) in pediatric patients attending a university-based infant oral health clinic. Key findings indicate that while race initially appeared to influence ECC risk, socioeconomic factors like parental health literacy and SES played a more substantial role after adjustment for other variables.

We also found that early detection, dental checkups, and preventive oral health education are critical in reducing ECC risk. Factors like frequent snacking and bottle use at night were associated with an increased risk of caries, suggesting the importance of dietary habits and feeding practices in ECC prevention.

Overall, the study’s findings emphasize the need for comprehensive preventive strategies that encompass both individual and community-level interventions. Early detection, effective prevention measures, and tailored oral health education are crucial to combat ECC, especially in underserved populations. Future research should explore the complex interactions between risk factors and their cumulative impact on ECC, which can lead to more effective and culturally sensitive intervention programs. By enhancing oral health promotion materials based on these insights, healthcare providers can contribute to more effective outreach and education efforts, ultimately reducing the prevalence and severity of ECC in pediatric populations.

## Data Availability

All data produced in the present study are available upon reasonable request to the authors

---

**Appendix 1:**
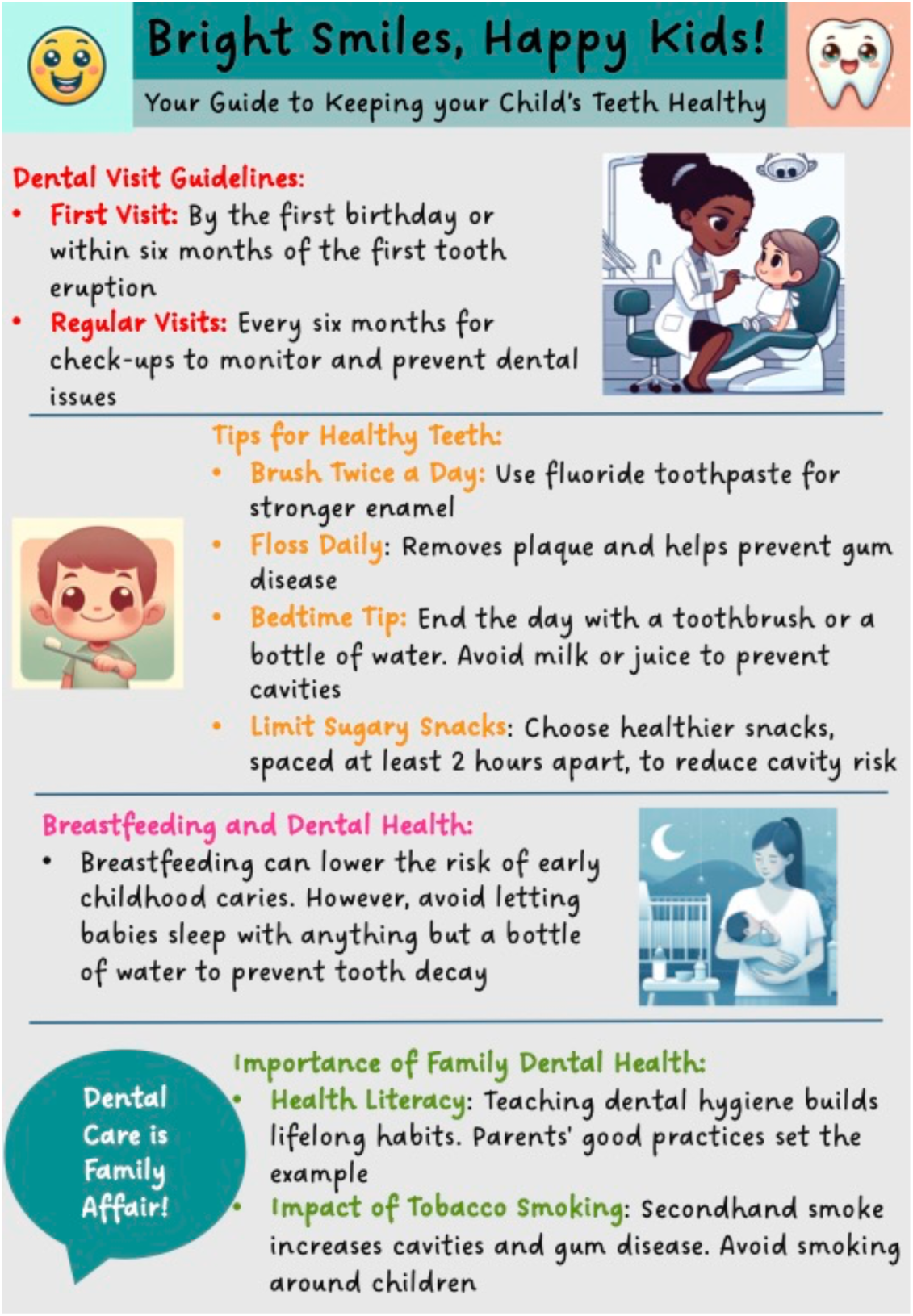
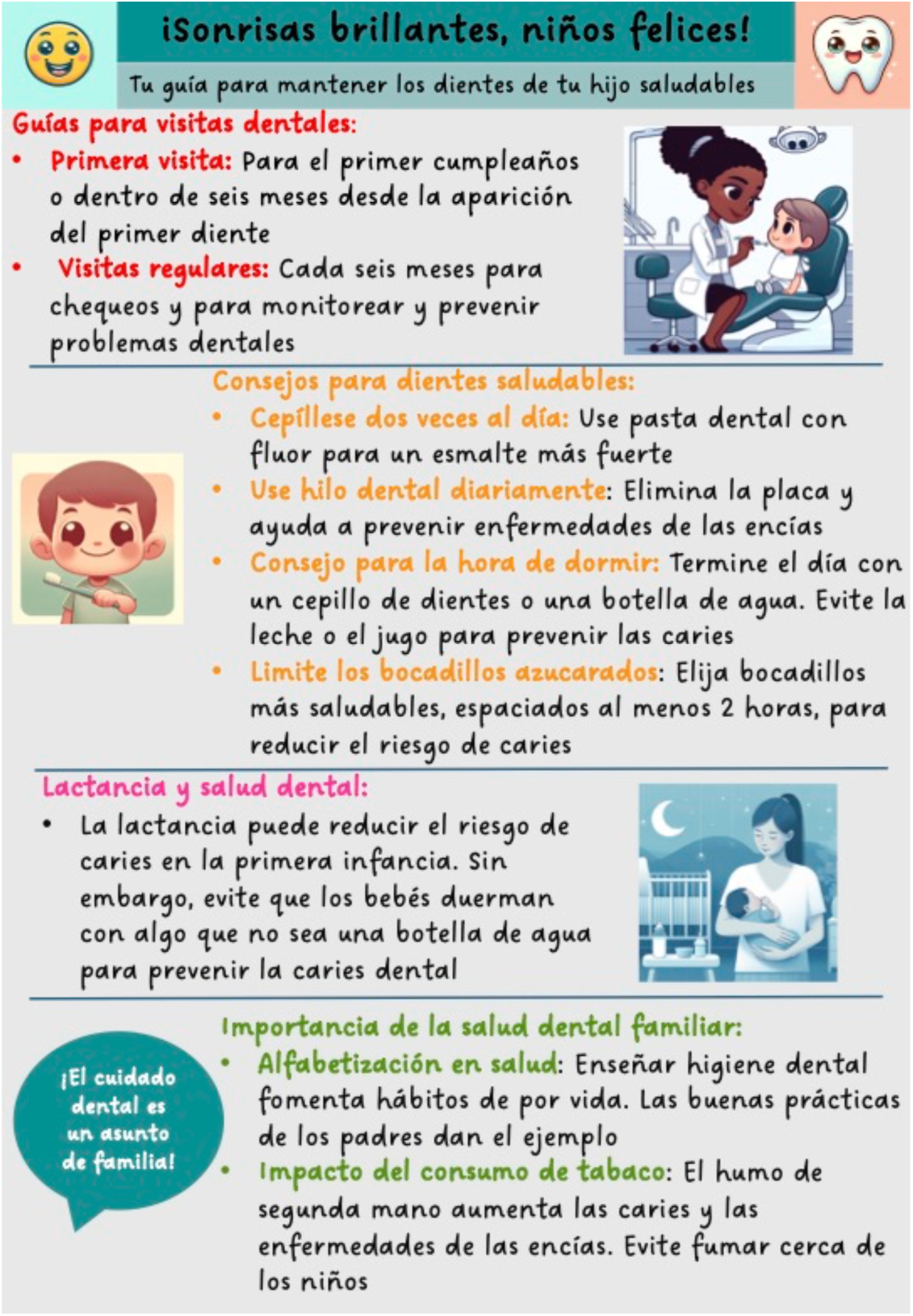
Promotional Material for Oral Health Education and Feedback Questionnaire.

